# Enhancing Migraine Trigger Surprisal Predictions: A Bayesian Approach to Establishing Prospective Expectations

**DOI:** 10.1101/2025.05.03.25326924

**Authors:** Dana P. Turner, Emily Caplis, Twinkle Patel, Timothy T. Houle

## Abstract

**Objective:** To extend the application of surprisal theory for predicting migraine attack risk by developing methods to estimate trigger variable likelihood in real time, under conditions of limited personal observation.

**Background:** Prior work has demonstrated that higher surprisal, a measure quantifying the unexpectedness of a trigger exposure, predicts headache onset over 12 to 24 hours. However, these analyses relied on retrospective expectations of trigger exposure formed after extended data collection. To operationalize surprisal prospectively, Bayesian methods could update expectations dynamically over time.

**Methods:** In a prospective daily diary study of individuals with migraine (N = 104), data were collected over 28 days, including stress, sleep, and exercise exposures. Bayesian models were applied to estimate daily expectations for each variable under uninformative and empirical priors derived from the sample. Stress was modeled using a hurdle-Gamma distribution, sleep using a rounded Normal distribution, and exercise using a Bernoulli distribution. Surprisal was calculated based on the predictive distribution at each time point and compared to static empirical surprisal values obtained after full data collection.

**Results:** Dynamic Bayesian surprisal values systematically differed from retrospective empirical estimates, particularly early in the observation period. Divergence was larger and more variable under uninformative priors but attenuated over time. Empirically informed priors produced more stable, lower-bias surprisal trajectories. Substantial individual variability was observed across exposure types, especially for exercise behavior.

**Conclusions:** Prospective surprisal modeling is feasible but highly sensitive to prior specification, especially in sparse data contexts (e.g., a binary exposure). Incorporating empirical or individually informed priors may improve early model calibration, though individual learning remains essential. These methods offer a foundation for real-time headache forecasting and dynamic modeling of brain-environment interactions.

## Introduction

Identifying the antecedents of migraine attacks remains a central challenge in headache science.^1^ While many individuals engage in self-monitoring of putative triggers, empirical associations between specific environmental or psychosocial exposures and subsequent migraine onset are often weak or inconsistent.^2–7^ In our previous work,^8^ we proposed a novel framework rooted in information theory, operationalizing *surprisal*, a measure that quantifies how unexpected or improbable an event is given recent prior experience, as a unifying construct for evaluating diverse antecedents or “triggers” of headache.^9^ Across multiple datasets, we found that higher surprisal values, representing more unexpected experiences, were associated with increased likelihood of imminent headache onset within 12–24 hours.^8,10,11^ These results suggest that it is not merely the presence or absence of a trigger that matters but the extent to which the trigger deviates from prior expectations.

This conceptualization is consistent with frameworks in computational neuroscience, particularly predictive coding models^12^ and the free energy principle.^13^ These theories posit that the brain functions as a hierarchical Bayesian inference engine, continuously generating predictions about incoming sensory input and updating those predictions in response to mismatches, or prediction errors, between expected and actual input.^14^ According to the free energy principle, the brain minimizes a variational bound on surprise (i.e., free energy) to maintain homeostasis and reduce uncertainty. Within this framework, surprisal may be interpreted as a measurable correlate of prediction error, reflecting the degree to which a given biopsychosocial event(s) challenges the brain’s internal model of the world.^15^ We posit that migraine attack risk may arise from the sum of such errors, reflecting the degree of threat originating from changes/challenges in the environment.

However, to render this theory actionable in applied settings on newly encountered individuals, an important issue must be addressed: how to estimate the likelihood of an event prior to extensive observation. Classical estimation of surprisal depends on well-sampled empirical distributions from which to base surprisal estimates. Yet, in ecological contexts, individuals regularly encounter novel or infrequently repeated events. Estimating the degree of surprisal for these events is akin to knowing how surprising an event is likely to be before having the benefit of weeks of data on which to base the calculation. In all our previous research into surprisal estimation,^8,10,11^ the information used to estimate surprisal was based on a retrospective expectation formed from the empirically observed distributions (i.e., we learned how surprising any exposure was after having observed it for weeks). For surprisal to serve as a practical tool for forecasting migraine attack risk or modeling real-time brain-environment interactions, it must be possible to approximate the expectedness of events under conditions of limited data. Accordingly, this study explores strategies for inferring event likelihood in low-data contexts, aiming to extend trigger-surprisal theory from a retrospective analytic tool to a forward-facing predictive framework.

## Methods

This study represents the third pre-specified primary analysis of a longitudinal dataset that has been detailed in two prior publications.^10,11^ Following approval by the local Institutional Review Board, data collection took place between April 2021 and December 2024 using a prospective design with daily diaries. Recruitment was conducted through multiple channels, including an institution-hosted online research platform, advertisements on public transportation, and community flyers. Interested individuals completed a telephone screening to assess eligibility. While full methodological details have been published previously,^10^ a brief summary is provided here for context. Eligible participants were adults aged 18 to 65 years with a diagnosis of migraine with or without aura as defined by the International Classification of Headache Disorders, 3rd Edition (ICHD-3)^16^ and who reported experiencing 4 to 14 headache days per month. Exclusion criteria included secondary headache disorders, chronic daily headache or medication overuse headache, recent changes in headache symptoms (within the past six weeks), insufficient English proficiency (below a sixth-grade level), unmanaged psychotic disorders, active substance dependence likely to affect headache activity or adherence to study procedures, and current or planned pregnancy during the course of the study.

Eligible participants completed an enrollment session, conducted either in person or virtually, during which informed consent was obtained via the electronic consent REDCap feature. Following consent, participants completed a battery of baseline questionnaires assessing demographics, headache characteristics, and migraine-related disability (e.g., Migraine Disability Assessment [MIDAS]^17^). They then received instruction in the at-home study procedures, which involved completing brief (5–10 minute) electronic diary entries twice daily, once each morning and evening, for a period of 28 days. All entries were submitted through REDCap.^18^ At study completion, participants attended a final session (in person or virtual) and completed a follow-up battery of questionnaires similar to those administered at enrollment. The twice-daily diary entries were designed to assess both exposure to a broad array of potential migraine triggers and headache activity. Details of the diary items and their variability have been published previously.^10^ Distinct sets of triggers were measured in the morning and evening diaries to reflect differing contextual influences. The morning (AM) diary focused on sleep patterns (duration, quality, awakenings, bedtime, and wake time), late-night eating, weather, and mood state (Profile of Mood States Short Form [POMS-SF]^19^). The evening (PM) diary also included the POMS-SF but emphasized common food and drink triggers, environmental exposures, caffeine and alcohol use, balance disturbances, missed meals, and daily stressors (Daily Stress Inventory [DSI]^20^), in addition to weather influences.

### Selection of Trigger Variables

Three biopsychosocial variables representing stress, sleep, and exercise were selected for dynamic surprisal modeling based on their theoretical relevance to headache risk,^3^ their practical feasibility for daily assessment, and their distinct measurement properties.^2^ Stress (i.e., daily hassles) was assessed using the intensity scale of the Daily Stress Inventory,^20^ yielding a unique continuous distribution with many zero counts (i.e., no stress) but also a positively skewed distribution. Sleep was measured as the self-reported number of hours slept the previous night, a continuous variable typically heaped at whole numbers due to rounding by the participants (see: ^21^). Exercise was captured as a self-reported binary indicator of whether a formal exercise session was completed each day (i.e., yes or no). These variables differ not only in their underlying distributions but also in the amount and type of information each provides for updating the expectations used for estimating surprisal. Stress ratings offer a wide but irregular range of possible values, sleep reports are constrained and discretized by natural rounding behavior, and exercise events provide sparse, dichotomous data. Consequently, each trigger variable introduces distinct inferential challenges for real-time probability estimation, particularly early in the observation period when limited data are available. Modeling all three allows for an examination of how surprisal behaves across variables with varying information, uncertainty structures, and learning dynamics, providing a broader examination of dynamic predictive frameworks in ecologically valid contexts.

### Prospectively Estimating Surprisal from Daily Observations

To quantify how unexpected an event is for each selected variable in real time, we implemented a Bayesian framework (see: ^22^) to dynamically estimate surprisal, defined as the negative logarithm of the predictive probability:

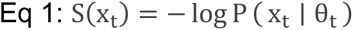

where x_t_ is the observed value on day t, and θ_t_ are the parameters of the predictive distribution at that time. Several methods for real-time probability estimation were considered, including kernel density estimation,^23^ Kalman filtering,^24^ and Bayesian structural time series.^25^ However, these approaches typically require extensive historical data or impose complex dynamics unsuitable for sparse, person-specific contexts. We therefore adopted a simple Bayesian updating framework optimized for limited daily observations.

Exercise, a binary variable (i.e., present or absent), was modeled using a Bernoulli likelihood with a Beta prior. After observing k exposures across t days, the posterior distribution was:

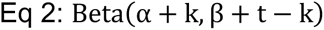

yielding a predictive probability of:

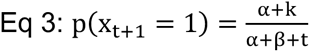

Sleep duration, a continuous but heaped variable, was modeled using a Normal likelihood with a conjugate Normal prior, rounded to the nearest integer hour to reflect observed reporting behavior. The posterior predictive distribution after t observations was:

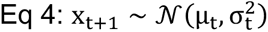

where μ_t_ is the updated posterior mean, and α_t2_^r^ eflects the combination of prior precision and accumulated evidence. To account for the common reporting behavior of rounding to the nearest whole hour, we computed the predictive probability of observing a specific integer value z by evaluating the cumulative density between adjacent half-integer bounds:

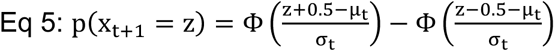

where Φ(⋅) denotes the cumulative distribution function (CDF) of the standard normal distribution. Thus, the probability of observing a sleep report of z hours corresponds to the probability mass between (z−0.5) and (z+0.5) under the day-specific predictive distribution. This approach preserves the probabilistic integrity of the continuous Normal model while honoring the discretized nature of the observed data. Surprisal for each sleep observation was then calculated by taking the negative logarithm of the corresponding rounded predictive probability.

Perceived stress was modeled using a hurdle-Gamma Bayesian framework. A Bernoulli model captured the probability of reporting zero stress, while a Gamma distribution characterized the magnitude of nonzero stress ratings. Given the lack of a true conjugate prior for the two-parameter Gamma distribution, we performed full Bayesian updating at each day by fitting a hierarchical hurdle-Gamma model using Hamiltonian Monte Carlo.^26^ Posterior distributions for the hurdle probability, Gamma shape, and Gamma rate parameters were estimated based on all accumulated observations through each day. The expected stress value on day t was computed as:

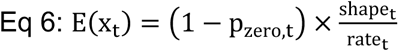

where p_zero,t_ represents the posterior hurdle probability of reporting zero stress.

### Prior Probability

Daily surprisal scores were computed under conditions using two distinct prior distributions. First, uninformative priors were specified to reflect minimal prior knowledge about the variable. For binary outcomes (e.g., exercise), we used a flat Beta prior:

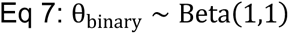

For sleep duration, a diffuse Normal prior was placed on the mean:

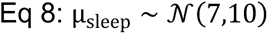

assuming a mean of 7 hours^27^ and very large prior variance (standard deviation ≈10 hours) to express broad uncertainty. For stress ratings, initial stress expectations were modeled with diffuse priors on the Gamma shape and rate parameters that matched our previous observations,^28^ each independently drawn from weakly informative Gamma distributions:

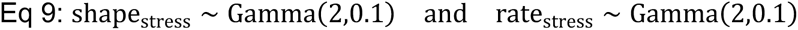

Second, empirical priors were derived individually using a leave-one-out strategy: for each participant, prior distributions were estimated from the empirical data of the remaining N−1 participants. The prior specification used the same distributions as in the uninformative priors, but for binary and sleep outcomes, empirical priors were centered on the observed means and variances of the sample excluding the target participant. For stress, empirical priors were based on maximum likelihood estimates of the Gamma distribution parameters fit to the leave-one-out sample. Posterior parameters were updated iteratively at each day for each individual, and surprisal values were calculated for every recorded observation, yielding 28 surprisal trajectories per participant per variable.

## Statistical Analyses

All statistical analyses were conducted using R version 4.4.1 and RStudio. All analyses were conducted with the available data, and no *a priori* statistical power calculations were conducted to guide sample size for this analysis. Descriptive statistics for participant characteristics were summarized using medians and interquartile ranges [25th, 75th] for continuous variables and frequencies and percentages for categorical variables. For dynamic modeling, Bayesian updating was applied iteratively to estimate daily expectations for each variable under both uninformative and empirically derived prior distributions. Daily expected values and surprisal scores were aggregated across participants to summarize group-level trajectories over the 28-day observation period.

For exercise, surprisal values were calculated separately for each participant, day, and prior type, then compared to static empirical estimates derived from each individual’s full data record. Mean differences between dynamic and static surprisal estimates were summarized at each timepoint, with 95% confidence intervals calculated based on the standard error of the mean. Visualization of distribution fits, expectation trajectories, and surprisal differences was performed using the ggplot2 package, with smoothing applied to trajectory plots where noted. All analyses were descriptive and exploratory with no formal hypothesis tests conducted.

## Results

Sample characteristics have been described previously.^10^ Briefly, the study included 109 individuals with migraine (median age = 35 years [IQR: 26–46]), 93.5% of whom were female and 83.5% of whom were White. Of these, 104 participants completed approximately 28 days of twice-daily electronic diaries, yielding 5,176 total entries. The sample had a median headache frequency of 8 days per month [IQR: 5–12], a median intensity of 7/10 [IQR: 5.5–8.0], and a median MIDAS score of 24 [IQR: 13–35.5], indicating moderate to severe migraine-related disability.

### Model Fit to Observed Distributions

Estimating surprisal in real time requires distribution-based expectations that reflect the empirical properties of each exposure variable. Stated differently, distributions that express the chances that someone encounters every level of the variable are required for the calculation. To allow this, observed distributions of daily stress, sleep, and exercise data were characterized by the proposed statistical distributions (Figure 1). Stress ratings exhibited a strong positive skew, with a high frequency of zero values. A hurdle-gamma model provided an appropriate fit, capturing both the probability of zero stress and the right-skewed distribution of nonzero values. The empirical cumulative distribution function (ECDF) aligned closely with the predicted cumulative distribution, indicating good model fit.

**Figure 1.**
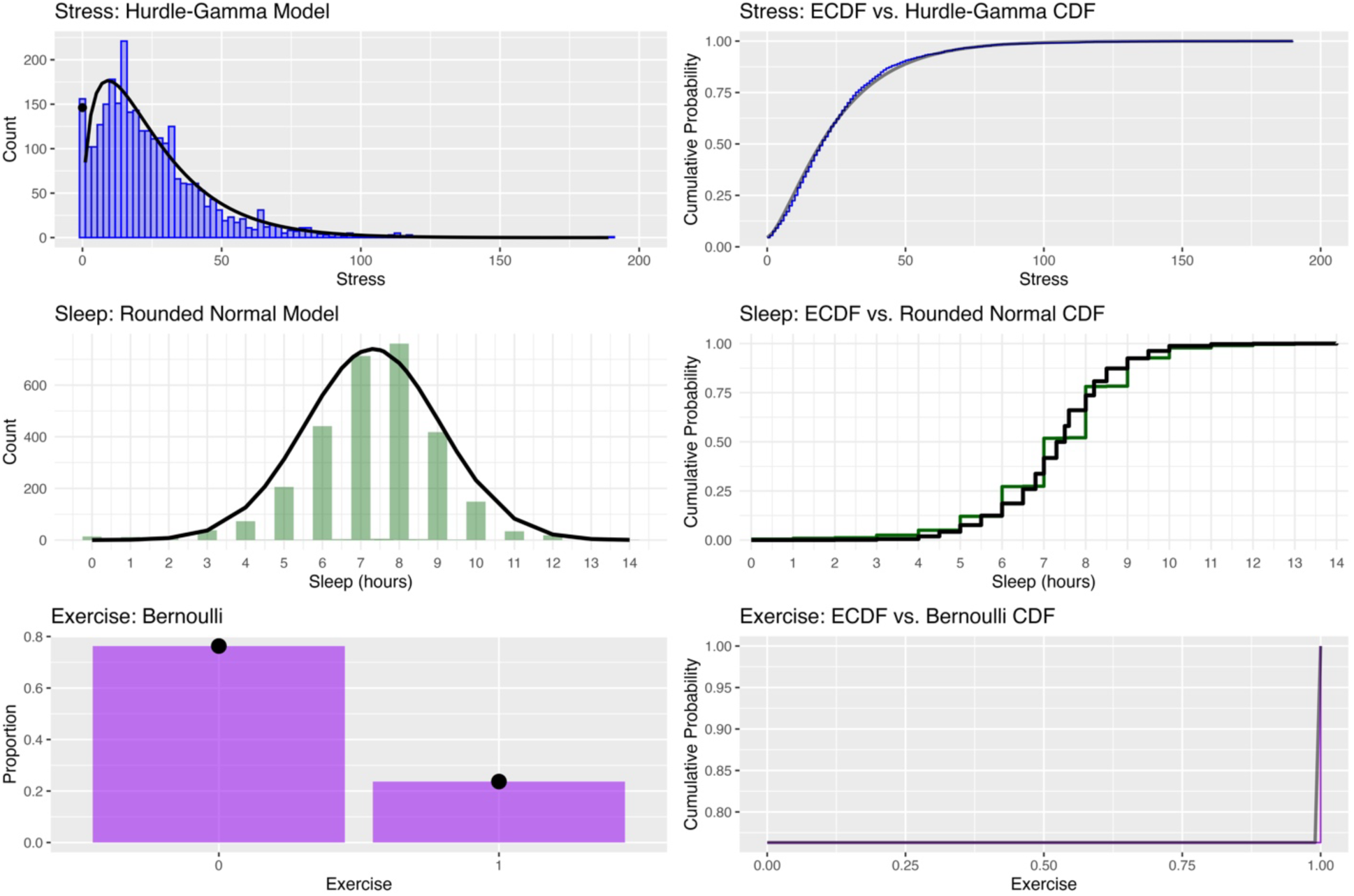
Observed and fitted distributions for stress, sleep, and exercise variables. Top left: Histogram and fitted hurdle-Gamma density for stress ratings, with the spike at zero reflecting the hurdle component. Top right: ECDF of stress ratings compared to the fitted hurdle-Gamma cumulative distribution. Middle left: Histogram of self-reported sleep duration overlaid with a rounded Normal density fit. Middle right: ECDF of sleep compared to the predictive cumulative distribution from the Normal model. Bottom left: Bar plot of exercise occurrence (0 = no, 1 = yes) with Bernoulli-predicted probabilities. Bottom right: ECDF of exercise compared to the Bernoulli cumulative distribution.

As expected, sleep duration was typically reported in whole-hour values. A rounded normal model was used to reflect this heaping. The fitted normal density provided an adequate approximation of the observed histogram, and the predicted cumulative distribution closely followed the ECDF. Finally, daily exercise, recorded as a binary variable (0 = no exercise, 1 = exercised), followed a Bernoulli distribution with a group-level mean near 0.25 (i.e., across the sample, individuals exercised on 25% of the days).

### Evolution of Expected Values Over Time

To assess the influence of prior knowledge on expectation formation, we compared temporal trajectories of estimated values generated from uninformative versus empirical priors. Bayesian expectation trajectories were generated for each individual using both uninformative (i.e., assuming no knowledge about the distribution) and empirical priors (i.e., using information provided by other participants) across the 28-day study period (Figure 2). For stress, expectations under empirical priors began higher than those under uninformative priors, but the two trajectories gradually converged over time as individual observations accumulated. Individual variability in stress reporting was considerable, particularly under the empirical prior, where a subset of participants showed persistently elevated expected values.

**Figure 2.**
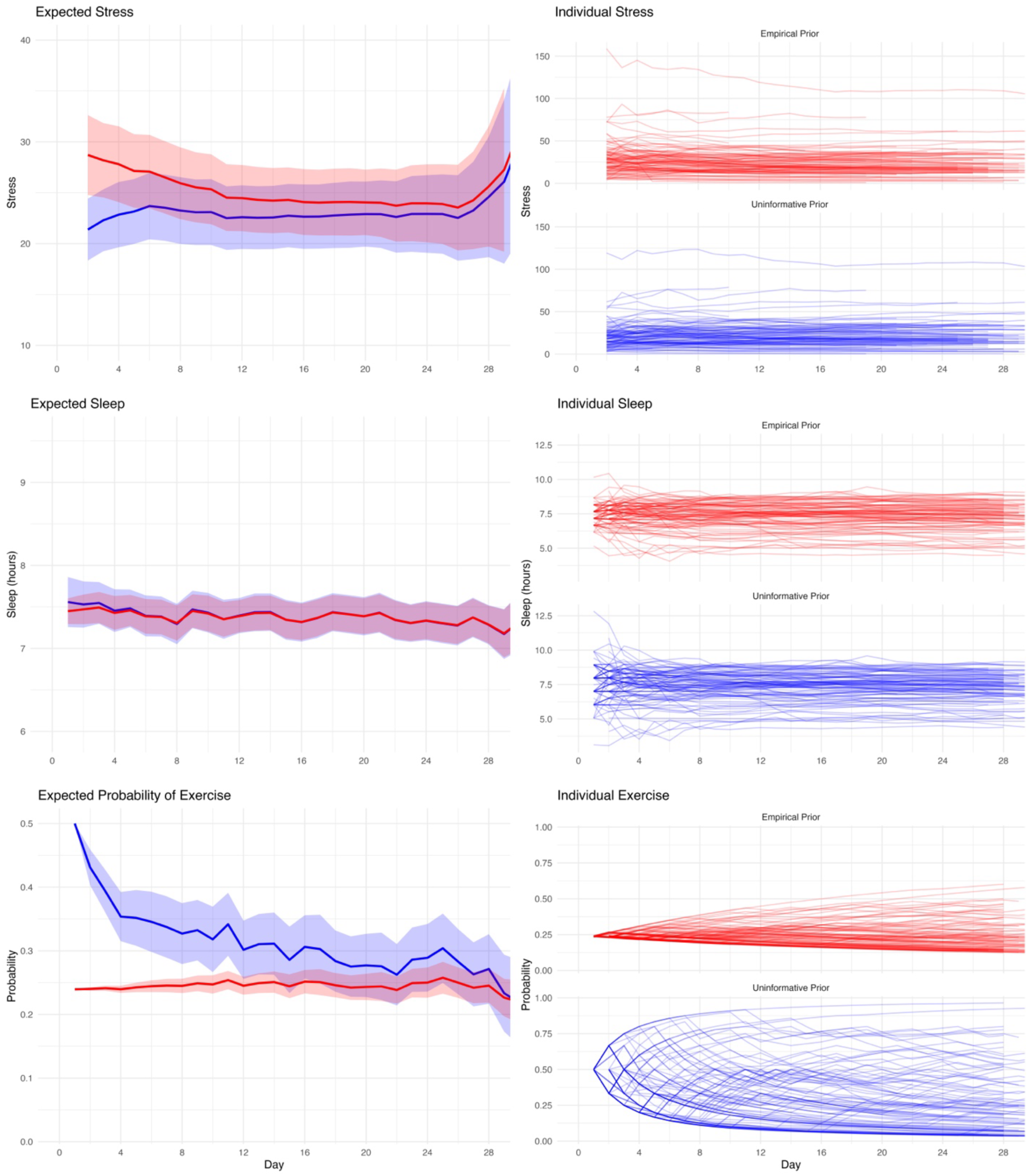
Evolution of expected values over time for stress, sleep, and exercise under uninformative (blue) and empirical (red) priors. Top left: Mean expected stress values across participants with 95% confidence intervals. Top right: Individual stress expectation trajectories. Middle left: Mean expected sleep duration over time. Middle right: Individual sleep expectation trajectories, showing intra-individual consistency. Bottom left: Mean expected probability of exercise over time. Bottom right: Individual exercise trajectories, highlighting wide between-person variability and prior-related differences in early estimates.

In contrast, sleep expectations remained relatively stable across time under both prior conditions. The empirical and uninformative priors produced similar group-level means, though the empirical prior provided some regularization early in the observation period, with greater between-person variability for the uninformative priors, as evidenced by more dispersed individual trajectories. This pattern supports the notion that individual sleep patterns vary across individuals but remain relatively consistent over time in the same individual.

For exercise, substantial divergence was observed between prior conditions. The uninformative prior initially assumed a relatively high probability of exercise (i.e., 50%), which decreased quickly with accumulating data. The empirical prior, by contrast, began with lower expectations that more closely reflected population averages and changed minimally over time. This difference in prior assumptions led to markedly different expectation curves, especially during the early observation period. Additionally, the very different pattern of individual trajectories supports strong individual differences across individuals that are only partially captured by the empirical prior, as the population average provides some information for expectation, but substantial individual differences remain that are learned during the observation period.

### Surprisal Differences by Prior Type

To evaluate the impact of prior specification on surprisal estimation, we compared dynamically updated Bayesian surprisal values to static empirical surprisal values, which were calculated using each individual’s full set of observed data and represent a reasonable benchmark for what could be learned with complete information. As shown in Figure 3 depicting exercise, the average surprisal difference (dynamic minus static) was negative for both prior types, indicating that dynamic surprisal values were generally lower than those based on a static empirical distribution. This difference was greatest early in the study period, reflecting greater uncertainty under limited data conditions.

**Figure 3.**
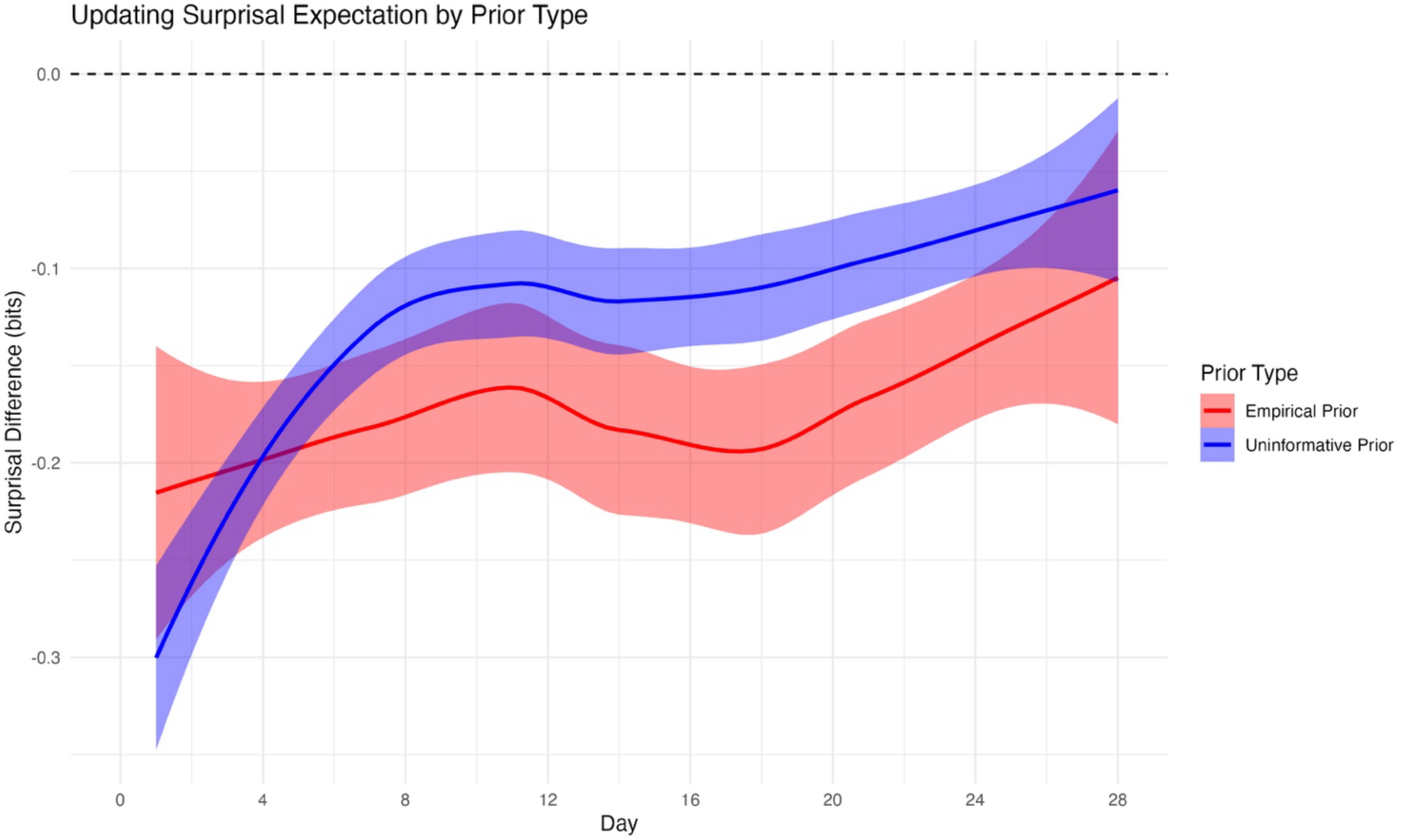
Mean difference between dynamic Bayesian surprisal estimates and static empirical surprisal values for exercise across the 28-day study period. Differences are plotted separately for empirical priors (red) and uninformative priors (blue), with shaded ribbons representing 95% confidence intervals. The dashed horizontal line at zero indicates perfect agreement. Dynamic surprisal estimates based on empirical priors exhibited smaller and more stable deviations from static estimates, whereas those based on uninformative priors showed greater early divergence that gradually diminished over time.

Notably, surprisal estimates based on the uninformative prior diverged more substantially from the static reference early on but began to converge toward the empirical estimates after approximately two weeks. In contrast, surprisal estimates under the empirical prior maintained a consistent bias with narrower confidence bands, suggesting more stable performance across time. These findings underscore the sensitivity of information-theoretic metrics to prior assumptions, particularly during the early phase of sequential data collection.

## Discussion

This study demonstrates a practical framework for estimating surprisal in real time by dynamically updating expectations using Bayesian inference. Building on prior work showing that surprisal predicts imminent migraine attacks,^8,11^ we sought to address a key limitation of prior methods: their reliance on retrospectively computed expectations from fully observed data. By incorporating both uninformative and empirically derived priors, we simulated the process of expectation formation under conditions of limited personal data, mirroring practical use cases in ecological forecasting and digital health applications.

The results illustrate how variable-specific measurement characteristics influence the learning dynamics of expectation and, by extension, surprisal. For example, in the case of stress, a highly skewed, semi-continuous variable, the empirical prior provided a strong initial approximation of expected values, which then adapted to individual idiosyncrasies over time. Conversely, for sleep, which followed a more stable but individualized pattern, expectations were relatively robust across both prior types, reflecting consistent intra-individual patterns. In contrast, exercise behavior exhibited substantial between-person variability and sparsity, making it especially sensitive to prior specification. These findings emphasize that the importance of inferring prior distributions for expectation formation is uneven across variable types, particularly in the early stages of data collection.

Importantly, we found that surprisal values based on dynamic Bayesian expectations may systematically diverge from hindsight-based estimates, particularly early in the observation period. This divergence was larger and more variable under uninformative priors, while empirically derived priors produced more stable trajectories, albeit with a modest and consistent bias relative to the fully informed benchmark. These findings highlight the importance of prior specification, not only for improving model stability and convergence, but also for enhancing the interpretability of applied surprisal values. In clinical and digital health contexts, where early inferences may inform behavioral guidance or therapeutic recommendations, the influence of prior choice is important.

One promising direction for improving early model calibration is in the use of individually informed priors. Such information could be obtained using baseline self-reports about typical sleep duration, exercise frequency, or stress levels. These personalized prior beliefs, elicited during study enrollment or digital onboarding, may offer a pragmatic middle ground between population-derived empirical priors and uninformed assumptions. Models based on these individualized assumptions could accelerate learning, reduce early surprisal volatility, and better reflect each individual’s expectations in the absence of long observation periods.

The study has several limitations. First, the analyses focused on modeling only three variables that differ in their statistical and behavioral properties. Generalization to other constructs or domains should be formally evaluated. Next, while the Bayesian models simulated real-time learning, all data were previously collected in a controlled observational context with structured diaries; performance may differ in passive sensing or naturalistic mobile data streams. Finally, the analysis ignored missing data in the diaries, as such information was not considered relevant for evaluating expectations over time. Yet, missing data that are conditional on a considered variable are almost certainly important in applied settings where participants may skip headache diaries when they are at particular risk for an attack.

In conclusion, these findings demonstrate that real-time surprisal modeling is feasible but highly sensitive to the choice of prior. Incorporating empirical or personalized priors may substantially improve model behavior in early phases of data collection, though individual learning remains essential for long-term calibration. Future work should explore how best to combine individual baseline information with adaptive prior structures.

## Data Availability

The data may be available upon reasonable request to the authors.

## Notes

Competing Interests: The authors declare no competing interests.

### Competing Interest Statement

The authors have declared no competing interest.

### Funding Statement

The research reported in this publication was supported by the National Institute of Neurological Disorders and Stroke of the National Institutes of Health under award number R01NS113823.

### Author Declarations

The Institutional Review Board of Mass General Brigham gave ethical approval for this work.

## References

1. Turner DP, Houle TT. Influences on headache trigger beliefs and perceptions. Cephalalgia. 2018;38:1545–53.

2. Turner DP. Assessing headache triggers: A practical guide for applied research and clinical management. Cham, Switzerland: Springer Nature. 2021.

3. Pellegrino ABW, Davis-Martin RE, Houle TT, Turner DP, Smitherman TA. Perceived triggers of primary headache disorders: A meta-analysis. Cephalalgia. 2018;38:1188–98.

4. Pavlovic JM, Buse DC, Sollars CM, Haut S, Lipton RB. Trigger factors and premonitory features of migraine attacks: Summary of studies. Headache. 2014;54:1670–9.

5. Lipton RB, Pavlovic JM, Haut SR, Grosberg BM, Buse DC. Methodological issues in studying trigger factors and premonitory features of migraine. Headache. 2014;54:1661–9.

6. Martin PR. Headache triggers: To avoid or not to avoid, that is the question. Psychol Health. 2000;15:801–9.

7. Martin PR. How do trigger factors acquire the capacity to precipitate headaches? Behav Res Ther. 2001;39:545–54.

8. Turner DP, Lebowitz AD, Chtay I, Houle TT. Headache triggers as surprise. Headache. 2019;59:495–508.

9. Turner DP, Caplis E, Bertsch J, Houle TT. Information theory and headache triggers. Headache. 2023;63:899–907.

10. Turner DP, Caplis E, Patel T, Houle TT. Development of a migraine trigger measurement system using surprisal. MedRxiv. 2025. medRxiv doi: 10.1101/2025.02.27.25322488

11. Turner DP, Patel T, Caplis E, Houle TT. Evaluating migraine trigger surprisal: Associations with migraine activity. MedRxiv. 2025. medRxiv doi: 10.1101/2025.04.23.25325821

12. Clark A. Whatever next? Predictive brains, situated agents, and the future of cognitive science. Behav Brain Sci. 2013;36:181–204.

13. Friston KJ. The free-energy principle: A unified brain theory? Nat Rev Neurosci. 2010;11:127– 38.

14. Spratling MW. A review of predictive coding algorithms. Brain Cogn. 2017;112:92–7.

15. Friston K, Da Costa L, Sajid N, Heins C, Ueltzhöffer K, Pavliotis GA, et al. The free energy principle made simpler but not too simple. Phys Rep. 2023;1024:1–29.

16. Olesen J. International Classification of Headache Disorders. Lancet Neurol. 2018;17:396–7.

17. Stewart WF, Lipton RB, Dowson AJ, Sawyer J. Development and testing of the Migraine Disability Assessment (MIDAS) Questionnaire to assess headache-related disability. Neurology. 2001;56:S20–8.

18. Harris PA, Taylor R, Thielke R, Payne J, Gonzalez N, Conde JG. Research electronic data capture (REDCap): A metadata-driven methodology and workflow process for providing translational research informatics support. J Biomed Inform. 2009;42:377–81.

19. Shacham S. A shortened version of the Profile of Mood States. J Pers Assess. 1983;47:305–6.

20. Brantley PJ, Waggoner CD, Jones GN, Rappaport NB. A Daily Stress Inventory: Development, reliability, and validity. J Behav Med. 1987;10:61–74.

21. Houle TT, Turner DP, Houle TA, Smitherman TA, Martin V, Penzien DB, et al. Rounding behavior in the reporting of headache frequency complicates headache chronification research. Headache. 2013;53:908–19.

22. Houle TT, Deng H, Tegeler CH, Turner DP. Continuous updating of individual headache forecasting models using Bayesian methods. Headache. 2021;61:1264–73.

23. Chen YC. A tutorial on kernel density estimation and recent advances [Internet]. arXiv [stat.ME]. 2017. Available from: 10.1080/24709360.2017.1396742

24. Harvey AC. Forecasting, structural time series models and the Kalman filter. Cambridge: Cambridge University Press. 1990.

25. Scott SL VHR. Predicting the present with Bayesian structural time series. Int J Math Model Numer Optim. 2014;5:4–23.

26. Carpenter B, Gelman A, Hoffman MD, Lee D, Goodrich B, Betancourt M, et al. Stan: A probabilistic programming language. J Stat Softw. 2017;76:1–32.

27. Steptoe A, Peacey V, Wardle J. Sleep duration and health in young adults. Arch Intern Med. 2006;166:1689–92.

28. Houle TT, Turner DP, Golding AN, Porter JAH, Martin VT, Penzien DB, et al. Forecasting individual headache attacks using perceived stress: Development of a multivariable prediction model for persons with episodic migraine. Headache. 2017;57:1041–50.

